# Dietary risk factors are prospectively associated with depression and anxiety over 3.5 years in mid-to-late adulthood: Findings from the English Longitudinal Study of Ageing

**DOI:** 10.1101/2025.10.30.25339197

**Authors:** Amelia McGuinness, Jasmine Cleminson, Adrienne O’Neil, Wolfgang Marx, Tetyana Rocks, Camille Lassale, Melissa Lane, Rebecca Orr, Gabriela Lugon, Felice Jacka, Deborah N Ashtree

## Abstract

**Objective:** To examine associations between intake of 21 food groups and nutrients and subsequent depression and anxiety in mid-to-late adulthood over 3.5 years.

**Design:** Prospective cohort study.

**Setting:** English Longitudinal Study of Ageing, a nationally representative study of community-dwelling older adults in England.

**Participants:** Adults (≥50 years) with complete dietary data, no prior depression or anxiety, and valid mental health outcomes at follow-up (N=2,754 depression; N=2,885 anxiety).

**Measurements:** Dietary intake was assessed in 2018-2019 using the Oxford WebQ 24-hour recall and categorized according to Global Burden of Disease dietary exposures and additional food groups and nutrients. Depression (CES-D-8≥3) and anxiety (GAD-7≥10) were assessed in 2021-23. Poisson regression estimated adjusted risk ratios (aRR) for associations between dietary exposures and incident depression or anxiety. This analysis forms part of the GLAD project (DERR2-10.2196/65576).

**Results:** Higher intakes of wholegrains (aRR=0.96; 95%CI:0.92-0.99 per 75g), fiber (aRR=0.83; 95%CI:0.71-0.98 per 30g), and iron (aRR=0.91; 95%CI:0.83-0.99 per 9mg) were associated with lower depression risk. Higher iron (aRR=0.80; 95%CI:0.64-1.00 per 9mg) and trans-fat (aRR=0.51; 95%CI:0.28-0.93 per 2% energy) intakes were associated with lower anxiety risk. Higher sugar-sweetened beverage intake was associated with both higher depression (aRR=1.05; 95%CI:1.02-1.09 per 335g) and anxiety (aRR=1.10; 95%CI:1.07-1.14 per 335g) risks.

**Conclusions:** Specific dietary components were prospectively associated with depression and anxiety risk over 3.5 years in mid-to-late adulthood. Sugar-sweetened beverages showed consistent adverse associations, while wholegrains, fiber, iron, and trans-fats appeared protective. The inverse association between trans-fat intake and anxiety warrants further investigation.

**Highlights:**

1. What is the primary question addressed by this study? What are the prospective associations between the intakes of 21 food groups and nutrients — including the globally recognized and policy-relevant Global Burden of Disease (GBD)-defined dietary exposures — and the risk of incident depression and anxiety over 3.5 years in mid-to-late adulthood?
2. What is the main finding of this study? In a cohort of adults aged ≥50 years, higher intakes of wholegrains, fiber, and iron were prospectively associated with lower risks of subsequent depression, while higher intakes of iron and trans fats were associated with lower risks of subsequent anxiety. Higher intakes of sugar-sweetened beverages were consistently associated with higher risks of both depression and anxiety.
3. What is the meaning of the finding? Certain dietary components were associated with the risk of developing depression or anxiety in adults aged 50 and over. While higher intakes of wholegrains, fiber, and iron appeared beneficial, and sugar-sweetened beverages detrimental, an unexpected inverse association with trans fat highlights the need for further investigation into its role in mental health.

## Objective

Common mental disorders are leading contributors to the global disease burden across the ageing population^1^. Depressive and anxiety disorders consistently rank among the top causes of disability from middle adulthood through to older age^1^. In adults aged 50–74 years, the burden of disability from depressive disorders exceeds that of several common chronic physical conditions, including colorectal cancer, osteoarthritis, and hypertensive heart disease^2^. Despite this sustained burden, poor mental health in mid-to-later life remains critically under-addressed. Even with the availability of effective and cost-effective treatments, low detection rates and poor coverage mean that only a minority of individuals with depressive disorders receive minimally adequate care^3,4^. These patterns underscore the urgent need to identify modifiable risk factors and strengthen preventive strategies across the ageing continuum.

Midlife and later adulthood are characterized by complex biopsychosocial transitions that can significantly increase vulnerability to mental disorders. These transitions encompass hormonal shifts, particularly menopause for women^5^, and significant life events including changes in income and occupational status due to retirement^6^, increased and often complex caregiving responsibilities^7^, and experiences of bereavement^8^. Declining physical and cognitive function^9,10^ as well as increased feelings of loneliness and isolation^11-13^, further elevate the risk of poor mental health in ageing populations. With projections indicating that one in six adults globally will be aged 60 or older by 2030^14^, identifying robust and modifiable risk factors to safeguard mental health throughout the second half of life is an essential public health imperative.

Diet is a modifiable risk factor significantly impacting mental health. Those in mid-to-late adulthood are particularly vulnerable to poorer dietary habits due to distinct nutritional challenges^15^. These include age-related changes in appetite, taste, smell, and metabolism, alongside medical comorbidities, changes to dentition, and polypharmacy, increasing vulnerability to malnutrition and micronutrient deficiencies^15-17^. Evidence from observational studies demonstrates that higher overall diet quality is associated with a reduced risk of depression in older populations. Specifically, adherence to ‘healthy’ dietary patterns like a Mediterranean-style diet, characterized by an abundance of plant foods, olive oil, fish, and minimally processed foods, commonly shows protective associations^18^. In contrast, ‘Western-style’ diets, characterized by highly processed foods, refined grains, and high sugar content, are linked to a higher risk of depression in later life^19^. Beyond broad patterns, research highlights the potential protective role of specific food groups, with high consumption of fruit and vegetables associated with reduced depression risk^19^. Furthermore, deficiencies in key nutrients such as long-chain omega-3 fatty acids, B vitamins (e.g., folate, B12), and vitamin D are increasingly implicated in the pathophysiology of depression in older adults^20^. In anxiety, although the evidence base is less extensive, findings are similar to depression; lower fruit and vegetable intake and higher saturated fat and added sugars are associated with higher risks for anxiety^21,22^. These diet-mental health connections are likely mediated through complex biological pathways, including the microbiota-gut-brain axis, modulation of systemic inflammatory processes, oxidative stress, and neurotrophic factor expression, all of which are critical for brain health and mood regulation in ageing^23,24^. As a fundamental and modifiable lifestyle behavior, diet offers a practical avenue to mitigate the prevalence and incidence of depression and anxiety in ageing populations.

While the existing literature has established a firm link between diet and mental health in older adults, several critical gaps remain. Most studies in this age group are cross-sectional, making it difficult to determine whether poor diet is a cause or a consequence of poor mental health. Many studies focus on prevalent symptoms rather than the onset of new disorders, limiting their ability to inform primary prevention strategies. Even in longitudinal research, the dietary exposures examined are often limited to a narrow selection of nutrients or food groups explored in isolation. Consequently, the relative importance of a comprehensive range of specific dietary components related to the development of mid-to-later life depression and anxiety is not yet clear. To address these limitations, the present study leveraged data from the English Longitudinal Study of Ageing (ELSA), a large, population-based cohort aged 50 years and over. By excluding participants with a history of depression or anxiety at baseline, we investigated the associations between dietary components and the incidence of these common mental disorders over a 3.5-year follow-up period. Critically, this is the first study to systematically apply the comprehensive Global Burden of Disease (GBD) framework, alongside other dietary exposures, to examine a wide array of specific food groups and nutrients as risk factors for the development of both depression and anxiety in an ageing population^25,26^. By doing so, this study aims to identify the most potent and modifiable dietary targets that may inform primary prevention strategies for common mental disorders in the second half of life.

## Methods

### Ethics

ELSA received ethical approval (February 2002) from the London Multi-Centre Research Ethics Committee (project number: MREC/01/2/91). The current project was approved by the Deakin University Human Research Ethics Committee (March 2024) for exemption from ethical review in accordance with the National Statement on Ethical Conduct in Human Research^27^ (project number: 2024-085). Methods were prospectively registered on Open Science Framework^28^ as part of the Global burden of disease Lifestyle And mental Disorders (GLAD) Taskforce^26^ which aims to generate global evidence regarding the association of GBD-defined dietary exposures with common mental disorders^26^.

### Setting

We used data from ELSA (accessed 18 March 2024)^29^. Details of ELSA have been published elsewhere^30^. Briefly, a representative sample of people aged ≥50 years was recruited from respondents of the Health Survey of England from 1998, 1999 and 2001. Data were collected via face-to-face, computer-assisted interviews or self-completed questionnaires. Participants were followed up biennially. Wave 9 (2018-2019; N=8,736) was the first year of dietary assessment^31^, considered the ‘baseline’ assessment for these analyses. Of the participants included at wave 9, N=4,329 completed the dietary questionnaire. For the purpose of our analysis, we excluded participants with no dietary data, aged <50 years, and with a history of depression or anxiety (Figure 1).

**Figure 1:**
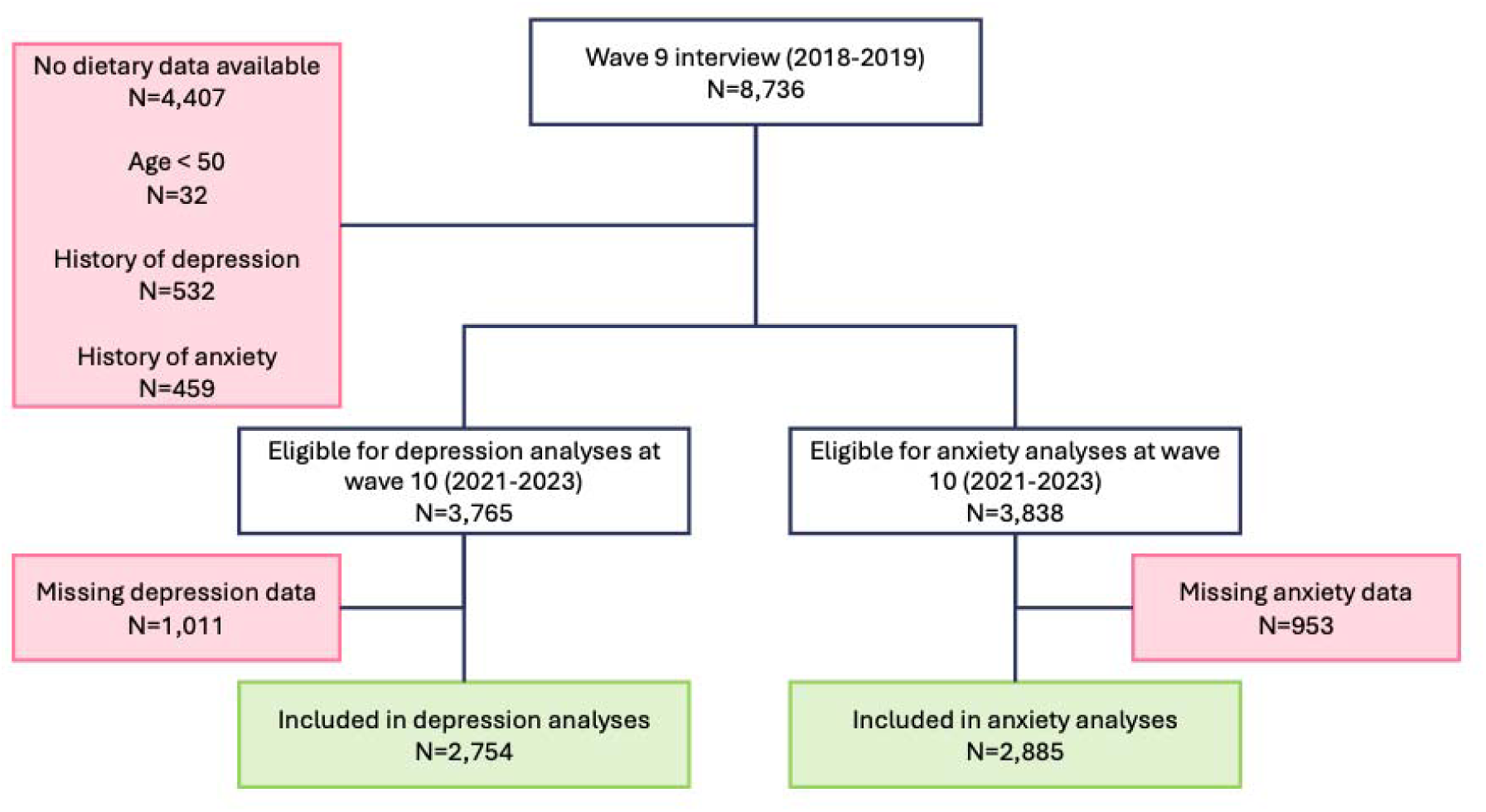
Participant flow chart from wave 9 of the English Longitudinal Study of Ageing, when diet was measured (considered the baseline for these analysis), to the final included sample at wave 10 for depression and anxiety.

### Exposures

Dietary intake was measured at wave 9 (2018-2019) on two non-consecutive days using the Oxford WebQ validated 24-hour dietary recall assessment tool^31^, as previously described^32^. Food items were recorded as portions per day, with nutrients estimated using data from the UK Nutrient Databank^33^. We used GBD-defined dietary exposures^34,35^, including omega-3, omega-6, polyunsaturated fat, trans fat, fruit, vegetables, legumes, wholegrains, nuts and seeds, milk, red meat, processed meat, sugar-sweetened beverages, fiber, calcium, and sodium. To match GBD units we multiplied portions per day by portion sizes for the UK^36^. In addition, we examined associations with saturated fat, monounsaturated fat, cholesterol, iron, and vitamin D, which may be particularly relevant in ageing due to their roles in maintaining neuronal membrane integrity, vascular and metabolic health, and neuroendocrine and immune function^37^.

The ELSA dataset did not include intakes for trans fat, sodium, omega-3, or omega-6. Therefore, we matched each ELSA food item to the food codes listed in the composition of foods dataset^33^. In cases where each ELSA food item was an aggregate of more than one food code, we disaggregated the food item into its components by using their corresponding proportions^36^. For example, chocolate biscuits included 25% from food code 7662 (cookies and biscuits with chocolate), 50% from food code 260 (digestive biscuits, half-coated in chocolate), and 25% from food code 10516 (American-style chocolate chip cookie)^36^. Sodium, omega-3, omega-6 and trans fat content of each ELSA food item was then obtained by multiplying the reported intake of that food item (in grams) by the sodium, omega-3, omega-6 and trans fat content from the food composition dataset, then summed to obtain each participant’s total daily sodium, omega-3 and trans fat intake.

Each dietary exposure was included as a continuous variable in the statistical models (% energy/day for omega-6, polyunsaturated fat, trans fat, monounsaturated fat, and saturated fat; µg/day for vitamin D; mg/day for omega-3, cholesterol, and iron; and g/day for all other variables). To aid with interpretation, results were transformed to be approximately equivalent to one recommended serving size for food groups or recommended daily intake (RDI) for nutrients, focusing on UK serving sizes or RDIs where possible^34,38-44^ (see Supplementary Material).

### Outcomes

Depressive symptoms were measured at wave 10 (2021-2023) using the eight-item version of the Center for Epidemiologic Studies Depression Scale (CES-D-8)^45^. Consistent with its established use in large, population-based cohort studies of ageing, including ELSA, a CES-D-8 score of three or higher was used to define the presence of likely cases of depression (henceforth “depression”) for our primary analyses^46-49^. Anxiety symptoms were measured at wave 10 using the seven-item Generalized Anxiety Disorder scale (GAD-7)^50^. A cut-off score ≥10 was used to identify likely cases of anxiety (henceforth “anxiety”). This is consistent with the original validation study wherein this threshold demonstrated a sensitivity of 89% and specificity of 82% for generalized anxiety disorder in clinical settings^50^.

Current or history of depression or anxiety was ascertained from waves 1-9 using the self-reported questions, “has a doctor ever told you that you had depression” and “has a doctor ever told you that you had anxiety”. Participants who reported a previous diagnosis of depression were excluded from the depression analyses (n=532), and those who reported a previous diagnosis of anxiety were excluded from the anxiety analyses (n=459).

### Covariates

We included self-reported demographic and general health information as covariates in these analyses, including sex (male/female), age (year), and education (categorical: college and above, A-levels, and O-levels and lower). To ensure consistency within the GLAD project, we used education as a proxy for socio-economic status^26^ and used Willett’s residual method to remove the potential confounding effect of energy intake^51^. Self-reported marital status (never married, formerly married (widowed, divorced or separated) and currently married/cohabiting); weight (kilograms); physical activity (metabolic equivalent of tasks minutes/day); lifetime smoking status (current, former, or never); alcohol consumption in the past 12 months (current, former, or never); and self-reported general health, mental ability and memory were additionally included as covariates in sensitivity models.

### Demographic Characteristics

Participant characteristics at baseline (wave 9) were reported as mean (SD) or median [Q1-Q3] for continuous variables and n (%) for categorical variables. To determine the percentages of participants with suboptimal dietary exposures, we used the GBD theoretical minimum risk exposure level (TMREL)^34,35^, or the mid-point of this where the TMREL was a range, for each dietary exposure. Participants were deemed to have suboptimal intake if they ate: <345g/day of fruit, <339g/day of vegetables, <105g/day of legumes, <185g/day of wholegrains, <21.5g/day of nuts and seeds, <310g/day (males) or <555g/day (females) of milk, >100g/day of red meat, >0g/day of processed meat, >0g/day of sugar-sweetened beverages, <23.5g/day of fiber, <0.79g/day (males) or <1.15g/day (females) of calcium, <565mg/day of omega-3, <9.5% daily energy from omega-6, <8% daily energy from polyunsaturated fatty acids, >0.55% daily energy from trans fat, and >3g/day of sodium.

### Statistical Analyses

A complete-case Poisson regression model with robust standard errors was used to estimate associations of each dietary exposure with risk of depression and anxiety, respectively. Participants with a history of depression and anxiety were excluded from their respective analyses. Two models were fitted for each dietary exposure and outcome pairing: 1) adjusted for energy using Willet’s residual method; and 2) adjusted for age, sex, education and energy (Willett’s residual method). We included subgroup analyses for: 1) males and females; and 2) age categories (50-64 years, 65-79 years, 80+ years). Additional sensitivity analyses included: 1) further adjusting for marital status, weight, physical activity, alcohol intake, smoking, general health, memory, and mental abilities; 2) determining the contribution of influential observations (observations with Cook’s distance>4/n-p were removed from the model)^52^; 3) determining the influence of extreme energy (participants with energy <1^st^ or >99^th^ percentile); and 4) imputing missing covariate and outcome data. All model assumptions were assessed prior to model fitting. Results are presented as adjusted risk ratios (aRR) with corresponding 95% confidence intervals (CIs). To account for multiple testing, p-values were adjusted using the Simes method (reported as q-values)^53^; however, interpretation was based on confidence intervals. All models were fitted in Stata 18.0.

## Results

### Participant characteristics

Of the N=2,754 participants with both valid baseline dietary data and follow-up depression data, N=1,370 met the threshold for depression after a mean follow-up period of 3.5±0.4 years. Of the N=2,885 participants with valid dietary and anxiety data, N=757 met the threshold for anxiety at follow up. For both depression and anxiety, a higher proportion of those who met the threshold were female, fewer had tertiary education, and a greater proportion reported poorer general health and mental abilities (Table 1).

**Table 1:** Baseline (wave 9) characteristics, including n (%), mean (SD) and median [Q1-Q3] dietary intake, of participants who responded to the dietary questionnaire and who were aged 50 and over at wave 9.

|  | No Depression at wave 10 (n=1,384) | Depression at wave 10 (n=1,370) | No anxiety at wave 10 (n=2,128) | Anxiety at wave 10 (n=757) |
| --- | --- | --- | --- | --- |
| Age (years) | 67.6 (7.3) | 68.8 (7.4) | 68.4 (7.3) | 67.8 (7.7) |
| Weight (kg) | 78.0 (14.8) | 78.0 (15.5) | 78.7 (15.3) | 76.1 (14.8) |
| Sex (% female) | 649 (46.9%) | 814 (59.4%) | 1070 (50.3%) | 471 (62.2%) |
| Education |  |  |  |  |
| College and above | 714 (51.6%) | 613 (44.7%) | 1065 (50.0%) | 328 (43.3%) |
| A-levels | 175 (12.6%) | 171 (12.5%) | 246 (11.6%) | 108 (14.3%) |
| O-levels or lower | 495 (35.8%) | 586 (42.8%) | 817 (38.4%) | 321 (42.4%) |
| Marital status |  |  |  |  |
| Never married | 55 (4.0%) | 57 (4.2%) | 79 (3.7%) | 40 (5.3%) |
| Formerly married | 193 (13.9%) | 240 (17.5%) | 338 (15.9%) | 123 (16.2%) |
| Currently married/cohabiting | 1136 (82.1%) | 1073 (78.3%) | 1711 (80.4%) | 594 (78.5%) |
| Lifetime smoking status |  |  |  |  |
| Never Smoked | 1361 (98.3%) | 1355 (98.9%) | 2098 (98.6%) | 746 (98.5%) |
| Former Smoker | 1 (0.1%) | 0 (0.0%) | 1 (0.0%) | 0 (0.0%) |
| Current Smoker | 1 (0.1%) | 0 (0.0%) | 2 (0.1%) | 0 (0.0%) |
| Not reported | 21 (1.5%) | 15 (1.1%) | 27 (1.3%) | 11 (1.5%) |
| Alcohol consumption (last 12 months) |  |  |  |  |
| Never drank | 144 (10.4%) | 181 (13.2%) | 237 (11.1%) | 104 (13.7%) |
| Former drinker | 246 (17.8%) | 282 (20.6%) | 399 (18.8%) | 160 (21.1%) |
| Current drinker | 994 (71.8%) | 907 (66.2%) | 1492 (70.1%) | 493 (65.1%) |
| Self-reported general health |  |  |  |  |
| Excellent | 276 (19.9%) | 140 (10.2%) | 366 (17.2%) | 72 (9.5%) |
| Very good | 536 (38.7%) | 475 (34.7%) | 810 (38.1%) | 231 (30.5%) |
| Good | 448 (32.4%) | 479 (35.0%) | 683 (32.1%) | 287 (37.9%) |
| Fair | 107 (7.7%) | 213 (15.5%) | 211 (9.9%) | 133 (17.6%) |
| Poor | 16 (1.2%) | 63 (4.6%) | 57 (2.7%) | 34 (4.5%) |
| Not reported | 1 (0.1%) | 0 (0.0%) | 1 (0.0%) | 0 (0.0%) |
| Self-rated memory |  |  |  |  |
| Excellent | 43 (3.1%) | 29 (2.1%) | 58 (2.7%) | 16 (2.1%) |
| Very good | 341 (24.6%) | 259 (18.9%) | 490 (23.0%) | 133 (17.6%) |
| Good | 677 (48.9%) | 661 (48.2%) | 1019 (47.9%) | 364 (48.1%) |
| Fair | 280 (20.2%) | 355 (25.9%) | 479 (22.5%) | 204 (26.9%) |
| Poor | 42 (3.0%) | 65 (4.7%) | 81 (3.8%) | 40 (5.3%) |
| Not reported | 1 (0.1%) | 1 (0.1%) | 1 (0.0%) | 0 (0.0%) |
| Self-rated mental abilities |  |  |  |  |
| Excellent | 181 (13.1%) | 117 (8.5%) | 242 (11.4%) | 67 (8.9%) |
| Very good | 532 (38.4%) | 498 (36.4%) | 846 (39.8%) | 228 (30.1%) |
| Good | 601 (43.4%) | 608 (44.4%) | 898 (42.2%) | 360 (47.6%) |
| Fair | 66 (4.8%) | 137 (10.0%) | 132 (6.2%) | 93 (12.3%) |
| Poor | 3 (0.2%) | 9 (0.7%) | 8 (0.4%) | 9 (1.2%) |
| Not reported | 1 (0.1%) | 1 (0.1%) | 2 (0.1%) | 0 (0.0%) |
| Energy kJ/day | 8627.7 (2521.4)<br>8425.2 [6863.5-10201.0] | 8449.1 (2554.6)<br>8255.7 [6738.4-9801.1] | 8532.3 (2501.8)<br>8339.3 [6765.7-9970.9] | 8450.2 (2698.8)<br>8183.2 [6717.0-9877.6] |
| Fruit intake (g/day) | 195.3 (171.4)<br>163.4 [70.0-275.5] | 198.2 (178.6)<br>160.9 [70.0-281.0] | 195.4 (178.3)<br>160.0 [66.0-275.5] | 200.3 (175.8)<br>164.0 [80.0-282.0] |
| Vegetable intake (g/day) | 207.1 (175.3)<br>172.8 [87.5-285.3] | 209.3 (189.5)<br>165.0 [77.5-292.5] | 205.0 (177.4)<br>166.9 [81.8-284.6] | 208.9 (193.2)<br>165.0 [76.2-291.8] |
| Legumes intake (g/day) | 35.4 (46.3)<br>19.8 [0.0-62.0] | 36.3 (48.4)<br>17.5 [0.0-59.0] | 35.4 (46.9)<br>17.5 [0.0-62.0] | 36.4 (48.5)<br>17.5 [0.0-52.5] |
| Wholegrain intake (g/day) | 70.7 (90.2)<br>36.0 [0.0-109.2] | 62.2 (77.1)<br>36.0 [0.0-101.8] | 67.4 (86.5)<br>36.0 [0.0-101.8] | 61.3 (76.2)<br>36.0 [0.0-98.0] |
| Nuts and seeds intake (g/day) | 7.3 (16.4)<br>0.0 [0.0-10.0] | 6.8 (16.2)<br>0.0 [0.0-0.0] | 7.0 (15.8)<br>0.0 [0.0-7.0] | 6.3 (16.7)<br>0.0 [0.0-0.0] |
| Milk intake (g/day) | 143.6 (129.4)<br>129.5 [64.8-194.2] | 148.4 (124.7)<br>129.5 [64.8-194.2] | 147.4 (129.9)<br>129.5 [64.8-194.2] | 140.2 (118.1)<br>129.5 [64.8-194.2] |
| Red meat intake (g/day) | 33.4 (47.2)<br>0.0 [0.0-60.0] | 34.9 (47.5)<br>0.0 [0.0-60.0] | 34.3 (47.7)<br>0.0 [0.0-60.0] | 31.3 (44.1)<br>0.0 [0.0-60.0] |
| Processed meat intake (g/day) | 16.5 (28.2)<br>0.0 [0.0-23.0] | 15.7 (27.2)<br>0.0 [0.0-23.0] | 16.2 (27.7)<br>0.0 [0.0-23.0] | 16.7 (28.3)<br>0.0 [0.0-23.0] |
| Sugar-sweetened beverages intake (g/day) | 226.3 (350.6)<br>46.2 [0.0-330.0] | 251.8 (379.9)<br>71.5 [0.0-377.5] | 224.4 (350.7)<br>0.0 [0.0-330.0] | 278.8 (408.3)<br>83.3 [0.0-455.0] |
| Fibre intake (g/day) | 16.2 (6.5)<br>15.5 [11.8-19.9] | 15.8 (6.8)<br>15.2 [11.0-19.4] | 15.9 (6.7)<br>15.2 [11.4-19.5] | 15.9 (6.7)<br>15.5 [11.4-19.5] |
| Calcium intake (g/day) | 0.9 (0.3)<br>0.9 [0.7-1.1] | 0.9 (0.3)<br>0.9 [0.7-1.1] | 0.9 (0.3)<br>0.9 [0.7-1.1] | 0.9 (0.3)<br>0.9 [0.7-1.1] |
| Omega-3 intake (mg/day) | 725.0 (686.1)<br>545.3 [210.4-1048.3] | 688.6 (633.9)<br>518.1 [190.8-1017.8] | 715.3 (674.4)<br>529.0 [198.8-1048.3] | 692.9 (626.8)<br>531.3 [201.2-1029.8] |
| Polyunsaturated fat intake (%energy/day) | 6.4 (2.3)<br>6.2 [4.8-7.8] | 6.3 (2.3)<br>6.1 [4.6-7.7] | 6.4 (2.3)<br>6.1 [4.7-7.7] | 6.3 (2.3)<br>6.1 [4.5-7.9] |
| Trans fat intake (%energy/day) | 0.3 (0.2)<br>0.3 [0.2-0.4] | 0.3 (0.2)<br>0.3 [0.2-0.4] | 0.3 (0.2)<br>0.3 [0.2-0.4] | 0.3 (0.2)<br>0.3 [0.2-0.4] |
| Sodium intake (g/day) | 1.3 (0.7) | 1.3 (0.7) | 1.3 (0.7) | 1.3 (0.7) |
|  | 1.2 [0.9-1.6] | 1.2 [0.9-1.6] | 1.2 [0.9-1.6] | 1.2 [0.9-1.6] |

### Dietary Intake

Across the sample, mean energy intake ranged from 8450 kJ (participants with depression or anxiety, respectively) to 8627.7 kJ (participants without depression), with approximately one-third of participants falling below estimated age-specific requirements for the United Kingdom^54^. A substantial proportion of participants did not meet the GBD-defined TMREL (Table 2). The majority had suboptimal intake of legumes (91.2%-92.3%), wholegrains (88.3%-91.9%), nuts and seeds (88.8%-90.8%), vegetables (80.3%-82.9%), fruit (83.4%-84.5%), milk (93.1%-95.9%), fiber (87.0%-87.7%), polyunsaturated fat (75.7%-77.4%) and omega-6 (98.9%-99.2%). Approximately half of the sample exceeded the TMREL for sugar-sweetened beverages (49.8%-55.2%). In contrast, excess intake of trans fat (11.2%-15.3%), red meat (10.3%-11.9%), and sodium (1.8%-2.2%) was relatively uncommon.

**Table 2:** Prevalence of suboptimal dietary intake, defined as not meeting GBD theoretical minimum risk exposure levels.

| GBD dietary risk | GBD TMREL | Dietary variables used in these analyses | No Depression at wave 10 (n=1,384) | Depression at wave 10 (n=1,370) | No anxiety at wave 10 (n=2,128) | Anxiety at wave 10 (n=757) |
| --- | --- | --- | --- | --- | --- | --- |
|  |  | Total energy intake (kJ/day) | 467 (33.7%) | 472 (34.5%) | 692 (32.5%) | 271 (35.8%) |
| Diet low in fruits | 340-350 g/day | Fruit (g/day) | 1170 (84.5%) | 1142 (83.4%) | 1790 (84.1%) | 635 (83.9%) |
| Diet low in vegetables | 306-372 g/day | Vegetables (g/day) | 1148 (82.9%) | 1100 (80.3%) | 1753 (82.4%) | 608 (80.3%) |
| Diet low in legumes | 100-110 g/day | Legumes (g/day) | 1277 (92.3%) | 1249 (91.2%) | 1954 (91.8%) | 694 (91.7%) |
| Diet low in wholegrains | 160-210 g/day | Wholegrains (g/day) | 1222 (88.3%) | 1259 (91.9%) | 1910 (89.8%) | 693 (91.5%) |
| Diet low in nuts and seeds | 19-24 g/day | Nuts and Seeds (g/day) | 1229 (88.8%) | 1233 (90.0%) | 1899 (89.2%) | 687 (90.8%) |
| Diet low in milk | 280-340 g/day (males); 500-610 g/day (females) | Milk (g/day) | 1289 (93.1%) | 1296 (94.6%) | 1981 (93.1%) | 726 (95.9%) |
| Diet high in red meat | 0-200 g/day | Red Meat (g/day) | 164 (11.8%) | 163 (11.9%) | 254 (11.9%) | 78 (10.3%) |
| Diet high in processed meat | 0 g/day | Processed Meat (g/day) | 599 (43.3%) | 564 (41.2%) | 908 (42.7%) | 329 (43.5%) |
| Diet high in sugar-sweetened beverages | 0 g/day | Sugar-Sweetened Beverages (g/day) | 698 (50.4%) | 730 (53.3%) | 1060 (49.8%) | 418 (55.2%) |
| Diet low in fibre | 22-25 g/day | Fibre (g/day) | 1214 (87.7%) | 1192 (87.0%) | 1858 (87.3%) | 664 (87.7%) |
| Diet low in calcium | 0.72-0.86 g/day (males); 1.1-1.2 g/day (females) | Calcium (mg/day) | 773 (55.9%) | 806 (58.8%) | 1208 (56.8%) | 451 (59.6%) |
| Diet low in seafood omega-3 fatty acids | 470-660 mg/day | Omega-3 (mg/day) | 717 (51.8%) | 734 (53.6%) | 1122 (52.7%) | 394 (52.0%) |
| Diet low in omega-6 polyunsaturated fatty acids | 9-10% daily energy | Omega-6 (% energy/day) | 1371 (99.1%) | 1356 (99.0%) | 2105 (98.9%) | 751 (99.2%) |
| Diet low in polyunsaturated fatty acids | 7-9% daily energy | Polyunsaturated Fat (% energy/day) | 1052 (76.0%) | 1061 (77.4%) | 1639 (77.0%) | 573 (75.7%) |
| Diet high in trans-fat | 0-1.1% daily energy | Trans-fat (% energy/day) | 199 (14.4%) | 193 (14.1%) | 326 (15.3%) | 85 (11.2%) |
| Diet high in sodium | 1-5 g/day | Sodium (g/day) | 31 (2.2%) | 24 (1.8%) | 39 (1.8%) | 17 (2.2%) |

\*For risks that are “diet low in”, the risk is calculated as number of people with intake less than the GBD recommended level, and for risks that are “diet high in”, the risk is calculated as number of people with intake higher than the GBD recommended level. Where the recommendations include a range, we used the midpoint (e.g. for fruit, the GBD recommendations are 340-350 grams, so we used 345 grams) ^The GBD do not have daily energy intake recommendations, so this is based on recommended energy intake for elderly people in the UK (8368 kJ for women and 10460 kJ for men)<sup>54</sup>. The risk here is presented as people eating less than the recommended daily energy intake.

### Depression

After adjusting for energy, age, sex, and education, higher intakes of wholegrains (aRR=0.96, 95%CI:0.92-0.99 per 75g serve), fiber (aRR=0.83; 95%CI:0.71-0.98 per RDI), and iron (aRR=0.91, 95%CI:0.83-0.99 per RDI) were associated with lower risks of depression. Conversely, higher intake of sugar-sweetened beverages was associated with higher risk of depression (aRR=1.05, 95%CI:1.02-1.09 per 335g serve). We observed no other associations between dietary intake and risk of depression in the full sample (Table 3).

**Table 3:** Association of dietary intake with risk of depression over 3.5 years.

| Exposure | Adjusted for energy<sup>^</sup> |  |  |  |  | Adjusted for energy<sup>^</sup>, age, sex and education |  |  |  |  |
| --- | --- | --- | --- | --- | --- | --- | --- | --- | --- | --- |
|  | RR | P | Q | LCI | UCI | RR | P | Q | LCI | UCI |
| Fruit (per 80g serve) | 1.01 | 0.429 | 0.674 | 0.99 | 1.02 | 1.00 | 0.773 | 0.854 | 0.98 | 1.01 |
| Vegetable (per 80g serve) | 1.01 | 0.472 | 0.708 | 0.99 | 1.02 | 1.00 | 0.722 | 0.824 | 0.98 | 1.01 |
| Legumes (per 150g serve) | 1.05 | 0.433 | 0.674 | 0.93 | 1.17 | 1.05 | 0.415 | 0.674 | 0.94 | 1.18 |
| Wholegrains (per 75g serve) | 0.96 | 0.018 | 0.126 | 0.92 | 0.99 | 0.96 | 0.024 | 0.126 | 0.92 | 0.99 |
| Nuts and Seeds (per 30g serve) | 0.98 | 0.679 | 0.824 | 0.92 | 1.06 | 0.99 | 0.701 | 0.824 | 0.92 | 1.06 |
| Milk (per 200g serve) | 1.04 | 0.241 | 0.481 | 0.98 | 1.10 | 1.04 | 0.226 | 0.481 | 0.98 | 1.10 |
| Red Meat (per 90g serve) | 1.04 | 0.252 | 0.481 | 0.97 | 1.12 | 1.04 | 0.267 | 0.488 | 0.97 | 1.12 |
| Processed Meat (per 70g serve) | 0.98 | 0.706 | 0.824 | 0.89 | 1.08 | 1.01 | 0.851 | 0.894 | 0.92 | 1.11 |
| Sugar-Sweetened Beverages (per 335g serve) | 1.04 | 0.022 | 0.126 | 1.01 | 1.07 | 1.05 | 0.003 | 0.042 | 1.02 | 1.09 |
| Fiber (per 30g RDI) | 0.94 | 0.003 | 0.042 | 0.91 | 0.98 | 0.83 | 0.023 | 0.126 | 0.71 | 0.98 |
| Polyunsaturated Fat (per 7% energy RDI) | 0.93 | 0.203 | 0.481 | 0.83 | 1.04 | 0.91 | 0.114 | 0.345 | 0.81 | 1.02 |
| Omega-3 (per 450mg RDI) | 1.00 | 0.017 | 0.126 | 1.00 | 1.00 | 1.00 | 0.207 | 0.481 | 1.00 | 1.00 |
| Omega-6 (per 2% energy RDI) | 0.90 | 0.519 | 0.752 | 0.66 | 1.23 | 0.90 | 0.539 | 0.755 | 0.66 | 1.24 |
| Calcium (per 700mg RDI) | 1.11 | 0.050 | 0.191 | 1.00 | 1.24 | 1.06 | 0.243 | 0.481 | 0.96 | 1.18 |
| Sodium (per 2g RDI) | 1.12 | 0.112 | 0.345 | 0.97 | 1.28 | 1.12 | 0.115 | 0.345 | 0.97 | 1.28 |
| Trans Fat (per 2% energy RDI) | 0.85 | 0.368 | 0.644 | 0.60 | 1.21 | 0.81 | 0.237 | 0.481 | 0.57 | 1.15 |
| Saturated Fat (per 11% energy RDI) | 0.98 | 0.726 | 0.824 | 0.89 | 1.09 | 0.97 | 0.572 | 0.775 | 0.87 | 1.08 |
| Monounsaturated Fat (per 13% energy RDI) | 0.90 | 0.038 | 0.160 | 0.81 | 0.99 | 0.89 | 0.186 | 0.481 | 0.76 | 1.05 |
| Cholesterol (per 300mg RDI) | 1.00 | 0.958 | 0.980 | 0.93 | 1.08 | 1.00 | 0.980 | 0.980 | 0.93 | 1.07 |
| Iron (per 9mg RDI) | 0.90 | 0.001 | 0.042 | 0.84 | 0.96 | 0.91 | 0.032 | 0.149 | 0.83 | 0.99 |
| Vitamin D (per 10µg RDI) | 0.99 | 0.797 | 0.858 | 0.96 | 1.03 | 0.99 | 0.595 | 0.781 | 0.95 | 1.03 |

<sup>^</sup>Energy adjustment was conducted using Willett's residual method and applied to dietary exposures reported in grams per day, but not those reported as % energy per day; RR=risk ratio; Q=Simes adjusted p-value; LCI=lower 95% confidence interval; UCI=upper 95% confidence interval; RDI=recommended daily intake

For males only, similar associations were observed for wholegrains, fiber, and sugar-sweetened beverages intakes with depression risk, however the association between iron and depression risk was no longer observed. In addition, higher monounsaturated fats were also associated with lower risk of depression, and higher sodium intake was associated with higher risk of depression (Supplementary Table 2).

For females only, similar associations were observed for iron and sugar-sweetened beverage intakes with depression risk, however the associations between wholegrains and fiber with depression risks were no longer observed. In addition, higher omega-3, omega-6, monounsaturated fat, and vitamin D intakes were all associated with lower risks of depression (Supplementary Table 2).

The associations between wholegrains, fiber, and sugar-sweetened beverage intakes with depression risk were observed across each age category. However, the association between higher iron intake and lower risk of depression was only observed for 50–64-year-olds. Additionally, higher red meat, processed meat, and sodium intakes were associated with higher risks of depression for participants aged 80 years and older (Supplementary Table 2). Results were similar in sensitivity models (Supplementary Table 3).

### Anxiety

After adjusting for energy, age, sex and education, higher intakes of trans fat (aRR=0.51, 95%CI:0.28-0.93 per RDI) and iron (aRR=0.80, 95%CI:0.64-1.00 per RDI) were associated with lower risks of anxiety. Conversely, higher sugar-sweetened beverage intake was associated with higher risk of anxiety (aRR=1.10, 95%CI:1.07-1.14 per 335g serve) (Table 4).

**Table 4:** Association of dietary intake with risk of anxiety over 3.5 years.

| Exposure | Adjusted for energy<sup>^</sup> |  |  |  |  | Adjusted for energy<sup>^</sup>, age, sex and education |  |  |  |  |
| --- | --- | --- | --- | --- | --- | --- | --- | --- | --- | --- |
|  | RR | P | Q | LCI | UCI | RR | P | Q | LCI | UCI |
| Fruit (per 80g serve) | 1.01 | 0.458 | 0.681 | 0.98 | 1.04 | 1.00 | 0.951 | 0.951 | 0.98 | 1.03 |
| Vegetable (per 80g serve) | 1.01 | 0.557 | 0.714 | 0.98 | 1.04 | 1.00 | 0.867 | 0.908 | 0.97 | 1.02 |
| Legumes (per 150g serve) | 1.05 | 0.604 | 0.725 | 0.87 | 1.27 | 1.06 | 0.578 | 0.714 | 0.88 | 1.28 |
| Wholegrains (per 75g serve) | 0.95 | 0.087 | 0.340 | 0.90 | 1.01 | 0.95 | 0.116 | 0.372 | 0.90 | 1.01 |
| Nuts and Seeds (per 30g serve) | 0.94 | 0.369 | 0.670 | 0.82 | 1.08 | 0.93 | 0.299 | 0.571 | 0.80 | 1.07 |
| Milk (per 200g serve) | 0.94 | 0.193 | 0.473 | 0.85 | 1.03 | 0.94 | 0.213 | 0.473 | 0.85 | 1.04 |
| Red Meat (per 90g serve) | 0.91 | 0.138 | 0.386 | 0.81 | 1.03 | 0.92 | 0.205 | 0.473 | 0.82 | 1.04 |
| Processed Meat (per 70g serve) | 1.05 | 0.560 | 0.714 | 0.90 | 1.21 | 1.07 | 0.399 | 0.670 | 0.92 | 1.25 |
| Sugar-Sweetened Beverages (per 335g serve) | 1.10 | 0.000 | 0.000 | 1.06 | 1.13 | 1.10 | 0.000 | 0.000 | 1.07 | 1.14 |
| Fiber (per 30g RDI) | 1.10 | 0.563 | 0.714 | 0.80 | 1.52 | 0.97 | 0.878 | 0.908 | 0.71 | 1.33 |
| Polyunsaturated Fat (per 7% energy RDI) | 0.93 | 0.470 | 0.681 | 0.77 | 1.12 | 0.89 | 0.227 | 0.477 | 0.74 | 1.07 |
| Omega-3 (per 450mg RDI) | 1.00 | 0.492 | 0.689 | 1.00 | 1.00 | 1.00 | 0.457 | 0.681 | 1.00 | 1.00 |
| Omega-6 (per 2% energy RDI) | 0.72 | 0.214 | 0.473 | 0.42 | 1.21 | 0.73 | 0.239 | 0.478 | 0.44 | 1.22 |
| Calcium (per 700mg RDI) | 1.20 | 0.038 | 0.266 | 1.01 | 1.42 | 1.16 | 0.084 | 0.340 | 0.98 | 1.38 |
| Sodium (per 2g RDI) | 0.97 | 0.813 | 0.899 | 0.78 | 1.21 | 0.98 | 0.886 | 0.908 | 0.78 | 1.23 |
| Trans Fat (per 2% energy RDI) | 0.47 | 0.013 | 0.182 | 0.26 | 0.85 | 0.51 | 0.027 | 0.252 | 0.28 | 0.93 |
| Saturated Fat (per 11% energy RDI) | 0.84 | 0.061 | 0.320 | 0.71 | 1.01 | 0.86 | 0.104 | 0.364 | 0.72 | 1.03 |
| Monounsaturated Fat (per 13% energy RDI) | 0.90 | 0.445 | 0.681 | 0.69 | 1.17 | 0.89 | 0.387 | 0.670 | 0.68 | 1.16 |
| Cholesterol (per 300mg RDI) | 1.02 | 0.753 | 0.855 | 0.91 | 1.14 | 1.02 | 0.705 | 0.822 | 0.91 | 1.15 |
| Iron (per 9mg RDI) | 0.78 | 0.030 | 0.252 | 0.63 | 0.98 | 0.80 | 0.045 | 0.270 | 0.64 | 1.00 |
| Vitamin D (per 10µg RDI) | 0.94 | 0.089 | 0.340 | 0.88 | 1.01 | 0.95 | 0.124 | 0.372 | 0.88 | 1.02 |

<sup>^</sup>Energy adjustment was conducted using Willett's residual method and applied to dietary exposures reported in grams per day, but not those reported as % energy per day; RR=risk ratio; Q=Simes adjusted p-value; LCI=lower 95% confidence interval; UCI=upper 95% confidence interval; RDI=recommended daily intake.

For males only, similar associations were observed for sugar-sweetened beverage intake with anxiety risk, however the associations between trans fat and iron intakes with anxiety risk were no longer observed (Supplementary Table 5).

For females only, similar associations were observed for iron and sugar-sweetened beverage intakes with anxiety risk, however the association between trans fat intake and anxiety risk was no longer observed. Higher omega-3 and vitamin D intakes were additionally associated with lower risk of anxiety (Supplementary Table 5).

The association between sugar-sweetened beverage intake and anxiety risk was observed across all age groups, however the association between iron intake and anxiety risk was only observed in those aged 50-64 years. The age group 65-79 years also had additional associations: higher saturated fat and vitamin D intakes were associated with lower risks of anxiety, and higher calcium intake was associated with higher risk of anxiety (Supplementary Table 5). Results were similar in sensitivity analyses (Supplementary Table 6).

## Conclusion

In this large, population-based cohort of adults aged 50 years and older, higher intakes of wholegrains, fiber, and iron were prospectively associated with lower risks of incident depression during the 3.5 years of follow-up, while higher intakes of iron and, surprisingly, trans fats were associated with lower risks of anxiety. Higher sugar-sweetened beverage intake was associated with higher risks of both depression and anxiety. Additionally, several associations differed by both sex and age bracket. Additionally, over 80% of participants fell short of recommended intakes for fruit, vegetables, legumes, wholegrains, seeds, milk, omega-6 polyunsaturated fatty acids, and fiber, and more than half exceeded recommendations for sugar-sweetened beverages. Together, these findings highlight that specific dietary components, and not just overall diet quality alone, are relevant to the risk of depression and anxiety in mid-to-late adulthood, and that sex- and age-related differences may shape these associations.

The inverse associations between wholegrains and fiber with risk of depression are consistent with findings from previous observational studies in older adults^55-58^. Intakes of these components may improve mental health by reducing systemic inflammation, modulating gut microbiota, and supporting neurogenesis and neurotransmitter function^24,59,60^, particularly at a period of vulnerability to neurodegeneration and neuroinflammation^61^. Fiber and wholegrains are rich in prebiotic substrates that promote gut-derived short-chain fatty acid production^62^, such as butyrate, which in turn, may influence brain function via the gut–brain axis^63^. These processes are increasingly implicated in the development of depression, and are of particular relevance for older adults, where age-related changes in immune function and brain plasticity may increase vulnerability to mood disorders^64,65^.

Counterintuitively, we observed an inverse association between trans fat intake and risk of anxiety. While industrially derived trans fats are well-established contributors to adverse cardiometabolic outcomes^66^, their relationship to mental health remains underexplored^67^. In this cohort, higher trans fat intake may reflect more ruminant-derived sources (e.g., dairy and meat) rather than highly processed snacks or fried foods^36^. This may serve as a proxy for more traditional, home-prepared dietary patterns such as the ‘meat and three veg’ model, which could signal greater nutritional adequacy. This association may also reflect unmeasured confounding, for example, individuals consuming traditional home-cooked meals might also benefit from stable eating routines, lower food insecurity, or more structured daily activities, which can independently support mental wellbeing^68^. However, in participants aged 80+ years, higher intakes of red and processed meats (common dietary sources of ruminant and industrial trans fats) and sodium were associated with increased risk of depression. These findings suggest that the relationship between certain dietary components and mental health may shift with age, underscoring the need for age-specific investigations into dietary risk and resilience factors.

Higher sugar-sweetened beverage intake was the only dietary factor consistently associated with greater risks of both depression and anxiety across models and subgroups, highlighting its potential importance as a modifiable prevention target. While sugar-sweetened beverages are often the focus of obesity and metabolic disease prevention^69^, their relationship with mental health is gaining recognition^70-72^, potentially through pathways involving blood glucose fluctuations, low-grade systemic inflammation, and gut–brain signaling^69,70,73^. In older adults, however, interpretation requires caution. Sugar-sweetened beverages may also include nutritionally fortified drinks such as oral nutritional supplements or meal replacements, commonly used to address undernutrition. While often containing added sugars and additives, they are serving a therapeutic function. As such, higher sugar-sweetened beverage intake may reflect not only discretionary consumption but also compensatory dietary strategies. This raises the possibility of residual confounding, where poorer underlying health status contributes both to supplement use and to increased vulnerability to depression and anxiety. Future research should aim to distinguish between discretionary and therapeutic sugar-sweetened beverage intake to more accurately assess mental health risks.

In females, greater intakes of iron, vitamin D, monounsaturated fats, and polyunsaturated omega-3 and omega-6 fatty acids were associated with lower risks of depression, and higher intakes of omega-3 and vitamin D were linked to lower anxiety risk. These findings may reflect the unique physiological and neuroendocrine changes that occur during mid-to-later life, such as menopause, which can increase susceptibility to mood disorders^74,75^. Iron plays a critical role in synthesizing monoamine neurotransmitters and deficiency (even without anemia) can impair mood regulation and increase fatigue^76^. This may be especially relevant for females, where estrogen influences dopaminergic function in the context of iron deficiency^76^. When stratified by age, the association between iron and depression was limited to those aged 50-64 years, aligning with the typical menopause transition period. Vitamin D, acting as a neurosteroid, influences brain function through anti-inflammatory effects, neurotrophic modulation, and serotonergic regulation^77^. As estrogen enhances vitamin D activation^78^, its decline in postmenopausal women may reduce vitamin D bioactivity, contributing to mood disturbances. Monounsaturated fats were associated with lower depression risk in both males and females, consistent with their established metabolic and neuroprotective benefits^79,80^. Protective associations for omega-3 and omega-6 fatty acids were observed only in females, which may reflect sex-specific differences in fatty acid metabolism, such as estrogen- or progesterone-enhanced PUFA synthesis and activity^81,82^, or behavioral adaptations during menopause, such as the adoption of heart-healthy diets rich in unsaturated fats (e.g., olive oil, fatty fish) to support cardiometabolic health.

Fewer dietary predictors of mental health emerged in males. As noted above, higher monounsaturated fat intake was linked to reduced depression risk, consistent with previous evidence that these fats support brain health^79,80^. Higher sodium intake, by contrast, was associated with greater risk of depression in men. While this may reflect physiological pathways, such as sodium’s influence on blood pressure, cardiovascular health, or HPA-axis regulation, measurement limitations warrant caution. Sodium intake from dietary questionnaires is often inaccurate^83^, and average levels in this cohort were lower than population norms^84^. Behavioral patterns may also play a role: men experiencing low mood may consume more processed or high-sodium foods. Still, emerging evidence suggests possible sex-specific neuroendocrine effects of sodium^85^, underscoring the need for further research into its role in male mental health.

This study has several strengths. We used data from a large, nationally representative, community-dwelling cohort of older adults living in England, with prospective ascertainment of mental health outcomes. The dietary variables were predominantly selected in line with those prioritized by the GBD, facilitating international comparison. Additional exposures were also selected, such as dietary lipids and micronutrients, to ensure a comprehensive analysis. Importantly, we excluded participants with a prior history of or prevalent depression or anxiety, strengthening the temporal inference between diet and incident mental health outcomes.

However, several limitations must be considered. First, dietary intake was assessed via two non-consecutive 24-hour recalls, which may not capture habitual intake or reflect seasonal variability. Although the Oxford WebQ is validated for use in large epidemiological studies, recall bias and misreporting remain possible. Second, while we adjusted for key sociodemographic confounders, residual confounding by unmeasured variables (e.g., chronic illness, psychosocial stressors) cannot be ruled out. Third, the outcome measures used (CES-D-8 and GAD-7) are screening tools, rather than clinical diagnostic tools, and while they are validated in older adults, misclassification is still possible and they may not capture sub-threshold cases. Finally, there was a relatively short timeframe between exposure and outcome in this study (3.5 years), which may limit the ability to observe long-term associations. Given this short follow-up window, the potential for reverse causation also cannot be excluded.

These findings have practical implications for public health. As the global population continues to age, the burden of common mental disorders in mid-to-later adulthood is likely to increase. Diet is a modifiable factor with population-wide exposure and strong public acceptability, making it a feasible target for mental health prevention and intervention efforts. Since small-scale improvements may be more acceptable and feasible than whole-of-diet changes^86^, improving specific food groups or nutrients may pose a suitable public health target to improving mental health in ageing populations. Our results suggest that promoting intake of wholegrains, fiber, health-associated fats, and micronutrients, while reducing sugar-sweetened beverage consumption, could offer a low-cost, scalable strategy to support mental health in mid-to-later adulthood. Given the observed sex differences, future research may benefit from sex-stratified analyses to guide personalized, sex-sensitive dietary strategies. Further research using repeated dietary assessments, intervention studies, and exploration of biological mechanisms will be critical to establish causality and optimize dietary recommendations for mental health in ageing societies.

## Data Availability

All data produced in the present study are available upon reasonable request to the authors

## Funding

AJM is funded through the National Health and Medical Research Council (NHMRC) supported CREDIT CRE, the Centre for Research Excellence for the Development of Innovative Therapies. JRC is funded through a NHMRC Medical Research Future Fund (MRFF) grant (MRF2033283). AON and DNA are supported by a NHMRC Emerging Leader 2 Fellowship (GNT2009295). FNJ is supported by a NHMRC Leader 1 Fellowship (GNT1194982). MML is supported by a Deakin University Postdoctoral Fellowship. RO is supported by a Deakin University Postgraduate Research Scholarship. WM is currently funded by an NHMRC Investigator Grant (GNT2008971). CL is supported by a Ramon y Cajal Fellowship (RYC2020-029599) funded by MICIU/AEI/10.13039/501100011033 and the European Social Fund “Invest in your future”. TR is supported by Roberts Family Foundation.

## Disclosures/Conflicts of Interest

AJM, JRC, RO, MML, TR, WM, AON, and DNA are members of the Food & Mood Centre, Deakin University, which has received research funding support from Be Fit Food, Bega Dairy and Drinks, and the a2 Milk Company, and philanthropic research funding support from the Waterloo Foundation, Wilson Foundation, the JTM Foundation, the Serp Hills Foundation, the Roberts Family Foundation, and the Fernwood Foundation. MML is a committee member and past secretary (2022–24) of the Melbourne Branch Committee of the Nutrition Society of Australia (unpaid) and has received travel funding support from the International Society for Nutritional Psychiatry Research, the Nutrition Society of Australia, the Australasian Society of Lifestyle Medicine, and the Gut Brain Congress and is an associate investigator for the MicroFit Study, an investigator-led randomized controlled trial exploring the effect of diets with varying levels of industrial processing on gut microbiome composition and partially funded by Be Fit Food (payment received by the Food & Mood Centre, Deakin University)

## Contributor Roles Taxonomy (CRediT)

Conceptualization: AON, FNJ, DNA, MML, RO

Data curation: DNA, CL

Formal analysis: DNA

Funding acquisition: AON

Investigation:

Methodology: AON, FNJ, DNA, MML, RO

Project administration: DNA

Resources: DNA, MML, RO

Software:

Supervision: AON, FNJ, DNA

Validation: AJM, DNA, RO

Visualization: AJM, DNA

Writing – original draft: AJM, DNA

Writing – review and editing: AJM, JRC, MML, CL, GL, WM, AO, RO, TR, FNJ, DNA

## Notes

### Author Declarations

The current project was approved by the Deakin University Human Research Ethics Committee (March 2024) for exemption from ethical review in accordance with the National Statement on Ethical Conduct in Human Research27 (project number: 2024-085).

